# COVID-19 Infection Forecasting based on Deep Learning in Iran

**DOI:** 10.1101/2020.05.16.20104182

**Authors:** Mehdi Azarafza, Mohammad Azarafza, Jafar Tanha

## Abstract

Since December 2019 coronavirus disease (COVID-19) is outbreak from China and infected more than 4,666,000 people and caused thousands of deaths. Unfortunately, the infection numbers and deaths are still increasing rapidly which has put the world on the catastrophic abyss edge. Application of artificial intelligence and spatiotemporal distribution techniques can play a key role to infection forecasting in national and province levels in many countries. As methodology, the presented study employs long short-term memory-based deep for time series forecasting, the confirmed cases in both national and province levels, in Iran. The data were collected from February 19, to March 22, 2020 in provincial level and from February 19, to May 13, 2020 in national level by nationally recognised sources. For justification, we use the recurrent neural network, seasonal autoregressive integrated moving average, Holt winter’s exponential smoothing, and moving averages approaches. Furthermore, the mean absolute error, mean squared error, and mean absolute percentage error metrics are used as evaluation factors with associate the trend analysis. The results of our experiments show that the LSTM model is performed better than the other methods on the collected COVID-19 dataset in Iran.

## 1. Introduction

Coronavirus disease 2019 is known as COVID-19 or 2019-nCoV which is an infectious disease. This virus leads to acute respiratory syndrome in many patients. It is also known as SARS-CoV-2 and is currently considered as international public health emergency [1]. The first alarm of COVID-19 disease was in December 2019 in Wuhan (capital of Hubei province) in China and has since spread globally were ongoing today as coronavirus pandemic [2,3]. The Secretary-General of the United Nations describes the COVID-19 disease as a global catastrophe 4 which directly threatens the human health, and affects the economic, social, environmental developments all around the world. This disease may change characteristics and epidemic mutates spatially and geographically based on global environmental changes which have not yet been sufficiently recognised [5]. In the other hand, by rapidity globalisation and concentration of population in cities, non-compliance of supply and demand for health services, shortcoming of medical protection, lack of expertise and mismanagements in developing countries has caused many difficulties for COVID-19 controlling and preventions. However, classifying and identifying sensitive areas in spatiotemporal distribution and transmissions of COVID-19, can be an important strategy to prevent the spread of the virus to other regions. Time series techniques based on deep learning can be proper approaches to forecast the behaviour of virus in order to achieve a correct and reliable prediction. Deep learning models for time series are used to predict the number of coronavirus infections over time. These models can forecast the near future and help to reduce the negative effects of the coronavirus.

In this article, we propose a deep-based approach for time series COVID-19 dataset to forecast the near future. We collect the data from February 19, to March 22, 2020 in provincial level and from February 19, to May 13, 2020 in national level by nationally recognised sources in Iran. Our experimental results show the effectiveness of the used approach in this study. The rest of the article is organised as follows. Section 2 introduces the methodology of the study were divided into the three parts included as data description, time series prediction procedure and Justification progress. Section 3 shows that the COVID-19 disease and infection development forecasting for Iran which classified in provincial and national levels. Section 4 is present the conclusion.

## 2. Materials and Methods

Since COVID-19 disease outbreak from the Wuhan city in China to the whole world, the scientists and specialists have different paths to recognising, analysing, controlling, improving, and cure this disease. In this circumstance, several epidemiological models are introduced by scholars which highly useful for estimating the dynamic transmission, targeting resources, and intervention strategies [6,7]. Therefore, the accurate forecasting of the future of COVID-19 requires the utilised of artificial intelligence techniques to overcome epidemiological models limitations. The presented study uses the long short term memory-based deep learning for time series forecasting of COVID-19 infectivity spatiotemporal distribution in Iran.

### 2.1 Data description

In this study, we collected COVID-19 confirmed data from February 19 (first infected) to March 22, 2020 in provincial level and February 19 to May 13, 2020 in national level. The provincial level data sources are from Iran ministry of health and medical education (http://behdasht.gov.ir) [8], Islamic republic news agency, IRNA (https://www.irna.ir) [9] and Iranian student’s news agency, ISNA (http://www.isna.ir) [10] reports. The national level data sources are from Iran ministry of health and medical education, statements of Dr. Kianush Jahanpur (spokesman for Iran ministry of health and medical education). Fig. 1 presents the infectivity rate as first day in 31 provinces of Iran. Furthermore, the Fig. 2 shows the infection variation during the February 19 to March 22, 2020 in each province. As seen in these figures, the Tehran, Isfahan, Mazandaran, Qom, Gilan, Alborz, Markazi, Khorasan Razavi, East-Azarbaijan, and Yazd provinces have the most affected and high risk until March 22, 2020. Prevalence of COVID-19 in Iran started on February 19, 2020 according to the report of the worldometers website [11]. Base on the report of the ministry of the health in Iran, by 2 reported COVID-19 cases, the virus is spread all over the country (on March 22, the confirmed cases reach to 21638). Fig. 3 shows the province level of the national distribution maps for new COVID-19 confirmed cases in Iran. It is noteworthy that after March 22, the province level infection data were not announced and this is the reason for collecting only until this date. It should be noted that the studied data are gathered by presented sources and conducted the long short-term memory-based deep learning (LSTM network) and several comparisons algorithms on the original data which has no interference on infections database in the studied time period.

**Figure 1.**
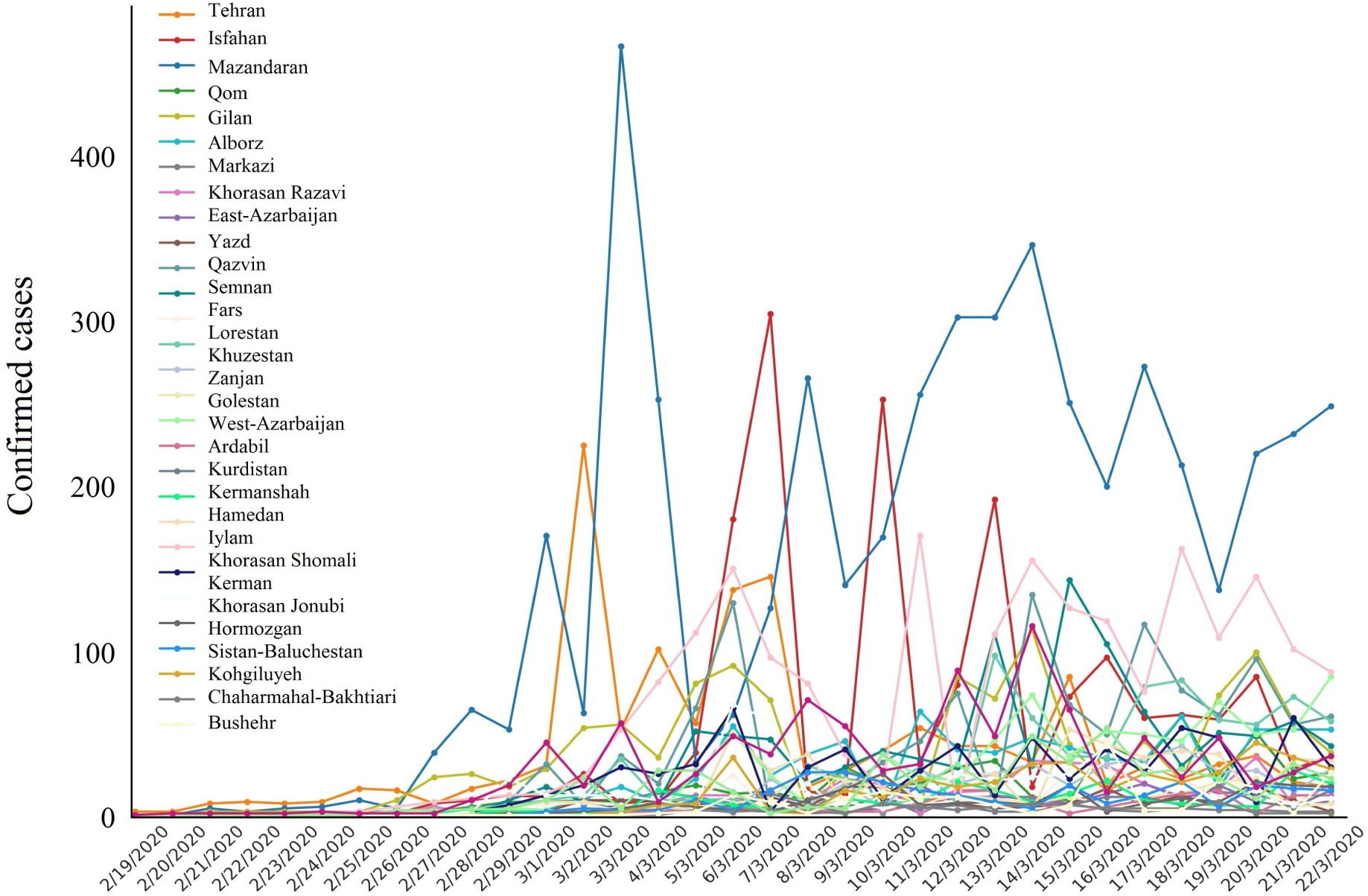
COVID-19 confirmed cases variation in Iran

**Figure 2.**
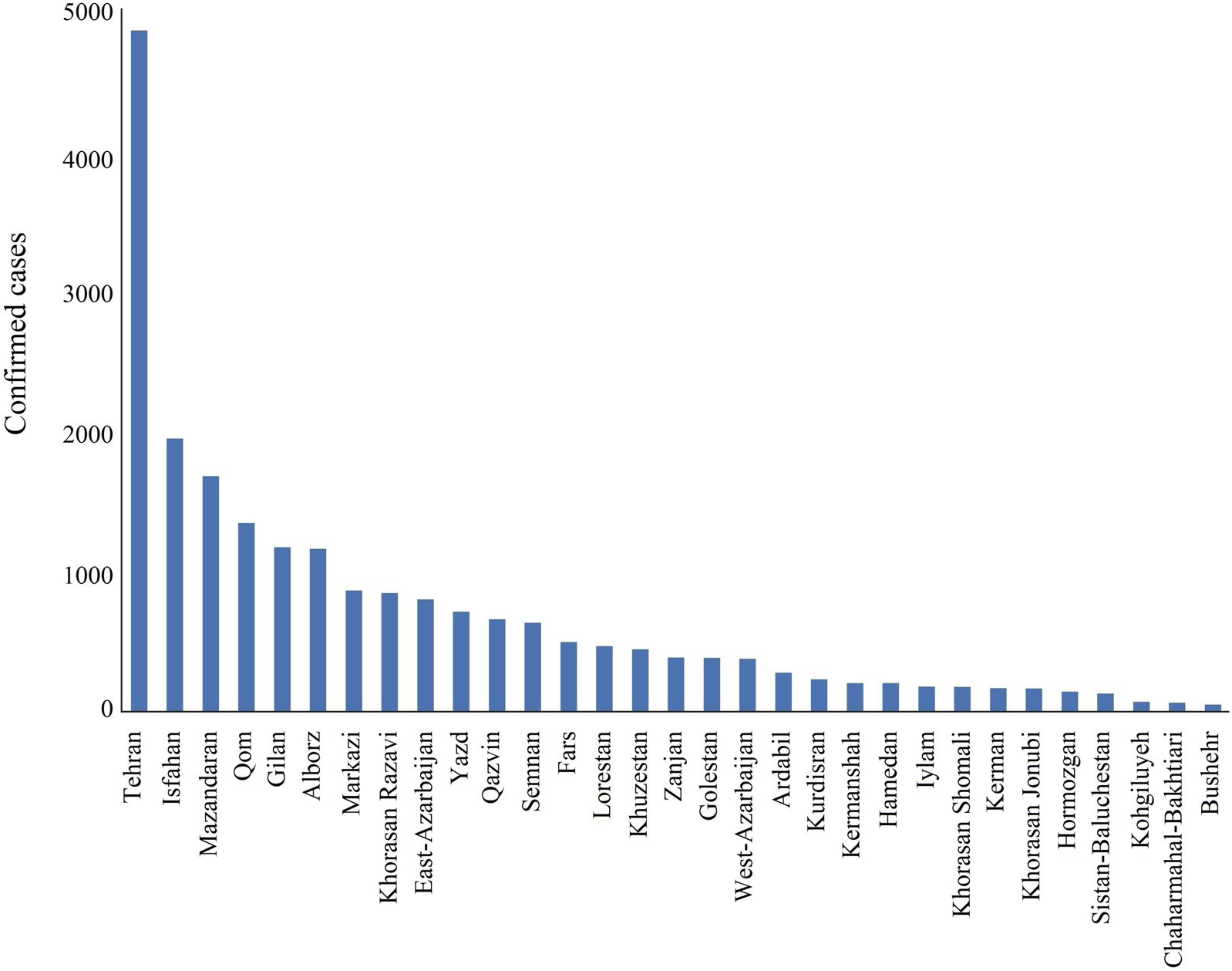
Cumulative results of confirmed cases in provincial level until March 22, 2020

**Figure 3.**
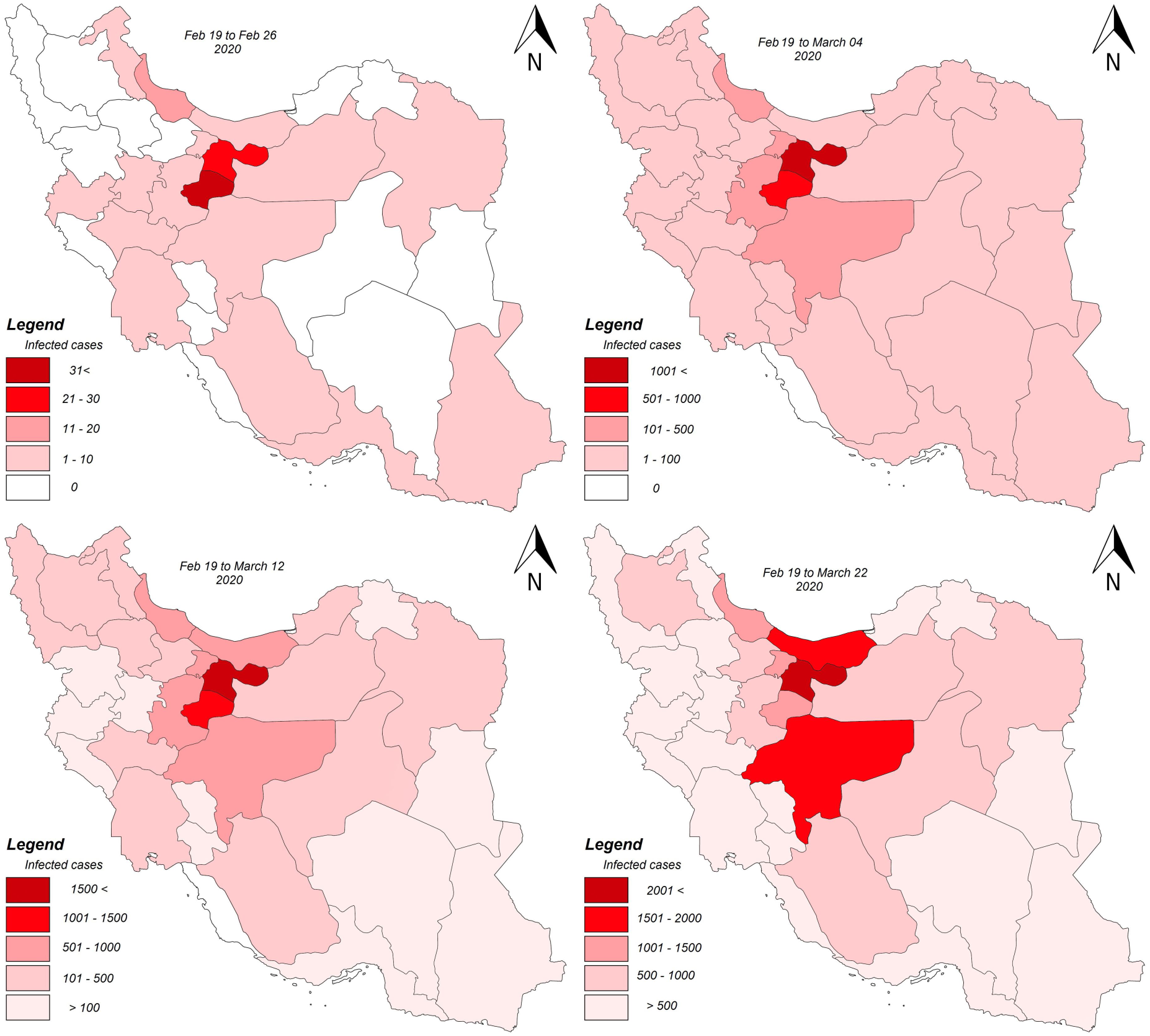
COVID-19 confirmed cases in Iran from 19 Feb 2019 to 22 March 2020 [8–10]

### 2.2 Time series forecasting procedure

The long short-term memory-based deep learning approach known as LSTM network used to the time series forecasting of infectious rate in Iran in two provinces and national levels from specific time period. Unlike traditional recurrent neural networks (RNN) where rewritten content at any time steps, the LSTM neural network is able to make decisions about retaining current memory through the specific gates. Intuitively, if the LSTM unit detects an important feature in the input sequence in the initial steps, it can easily transmit this information over a long distance. Therefore, it receives and maintains such long-term dependencies [12,13]. Time series forecasting can be found in several application domains, such as business, economics, inventory control, weather forecasting, and signal processing. By using LSTM’s time-related advantages for infection development forecasting, we are able to forecast temporal near future and contagious progress in Iran. In this regard, the provinces datasets (province level) are trained for 22 days and then tested for 12 days, all infection until 22 March in Iran increased to 21638 cases. Meanwhile, for entire country (national level) dataset is trained for 67 days and then tested for 19 days where covered the infection data from February 19 to May 13, 2020 (86 days). The LSTM neural network was implemented with Adam optimizer; the implementation was in TensorFlow (Python deep learning library).

### 2.3 Justification progress

In order to assess the capability of used technique, we used several forecasting methods to evaluate the results. We employed recurrent neural network (RNN), seasonal autoregressive integrated moving average (SARIMA), Holt winter's exponential smoothing (HWES), moving averages approaches in the comparisons. Some scholar used the forecast for the trend analysis of COVID-2019 pandemic development [14–21]. Thus, in this study RNN, SARIMA, HWES and Moving Average methods used as comparative procedures for LSTM. Meanwhile, the mean absolute error (MAE), mean squared error (MSE), and mean absolute percentage error (MAPE) criteria’s [22,23] are used as evaluation metrics.

## 3. Results and Discussion

In order to forecasting the time series two steps are considered for evaluating the COVID-19 pandemic development in Iran which are the province and national levels. In province level, we select 4 critical provinces, named Tehran, Isfahan, Mazandaran, and Qom, see Fig. 3. Based on the results of LSTM, we observed that high quick infections in near future for these provinces. As national level, the algorithm implemented to assess and forecast COVID-19 prevalence in the country at coming days. In the second experiment, we employ 33 days collected data for the provinces. In this dataset we consider 22 days as training and 12 days as test data. Figures 4 to 7 show the results of time series forecasting for Tehran, Isfahan, Mazandaran, and Qom provinces.

**Figure 4.**
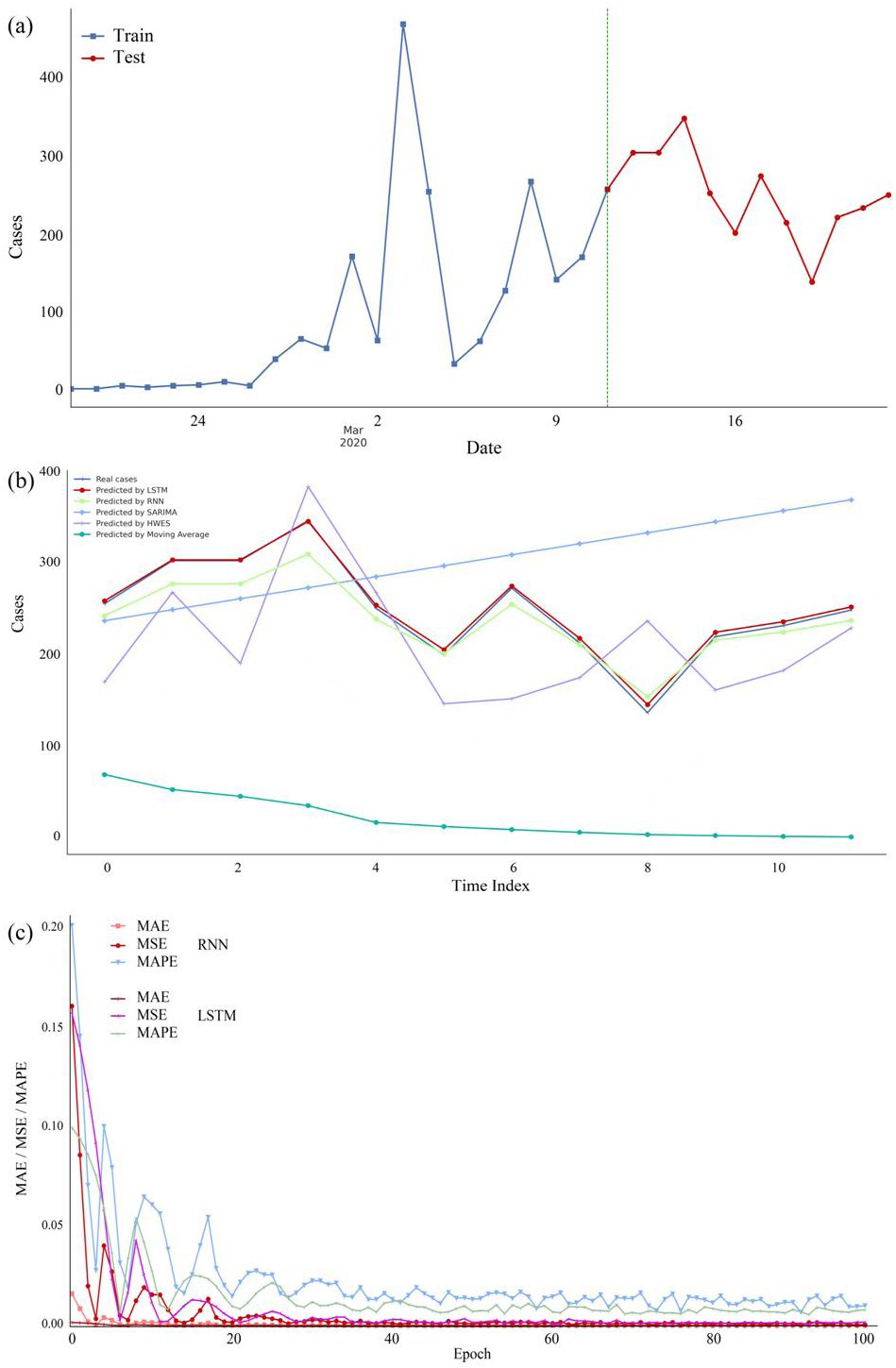
Infectious forecasting model for Tehran province based on deep learning: (a) Train and test days, (b) Forecasting and actual infected cases with different approaches (LSTM, RNN, SARIMA, HWES, and moving average), (c) Error models for RNN and LSTM

The Tehran city with about 22.7% of the total number of infections is the first place in Iran which has the largest confirmed cases until March 22, 2020. This statistic has shifted significantly between March 3 to 5 and March 10 to 13, 2020; but the infections number has fallen in recent days, see Fig. 4. The LSTM based forecasting is very close to real data and the model error rate is very low for all used metrics. After Tehran, Isfahan has a high number of confirmed infection cases with nearly about 9.1 percent on March 22, 2020. According to Fig. 5, the predicted data from the LSTM model for Isfahan is very close to reality. On March 5 to 8 and March 11 to 15 has significant shift in infections numbers in this province. The highest pollution level for Isfahan is on March 11 which is also the first peak day of comparison, see Fig. 5b. Mazandaran province had the third most risk-able provinces of Iran until March 22, 2020 with about 7.9% of confirmed infection cases as shown in Fig. 6. The highest incidence days in this province are on March 6, 7, and 13 with 180, 305, and 192 cases. Qom province was about 6.3% of all infection until reviewed date, as shown in Fig. 7. According to the results for these provinces the LSTM based algorithm has very high performance to forecast the infection rates.In national level, the prepared data (data sources mainly from Iran Ministry of Health and Medical Education [8]) which is gathered from February 19 to May 13, 2020 for entire country. Based on the collected 86 days’ data, 67 days are selected for training and 19 days for testing in the LSTM network. The total number of infected cases on May 13, 2020 was 112,725 which are obtained daily in this study. By evaluating and comparing to other methods, the proposed model has the highest proximity to reality in terms of forecasting. Furthermore, based on the error rate (MAE, MSE, MAPE), the proposed model has less error rate compare to RNN. Figure 8 shows the forecasting progress for the national level variation of the infection cases in Iran. As seen in this figure, the LSTM-based forecasting has higher accuracy. Meanwhile, figure 9 depicts the results of probable future in the next 21 days. According to this figure, we observe a descending course for infection development in Iran in coming two weeks.

**Figure 5.**
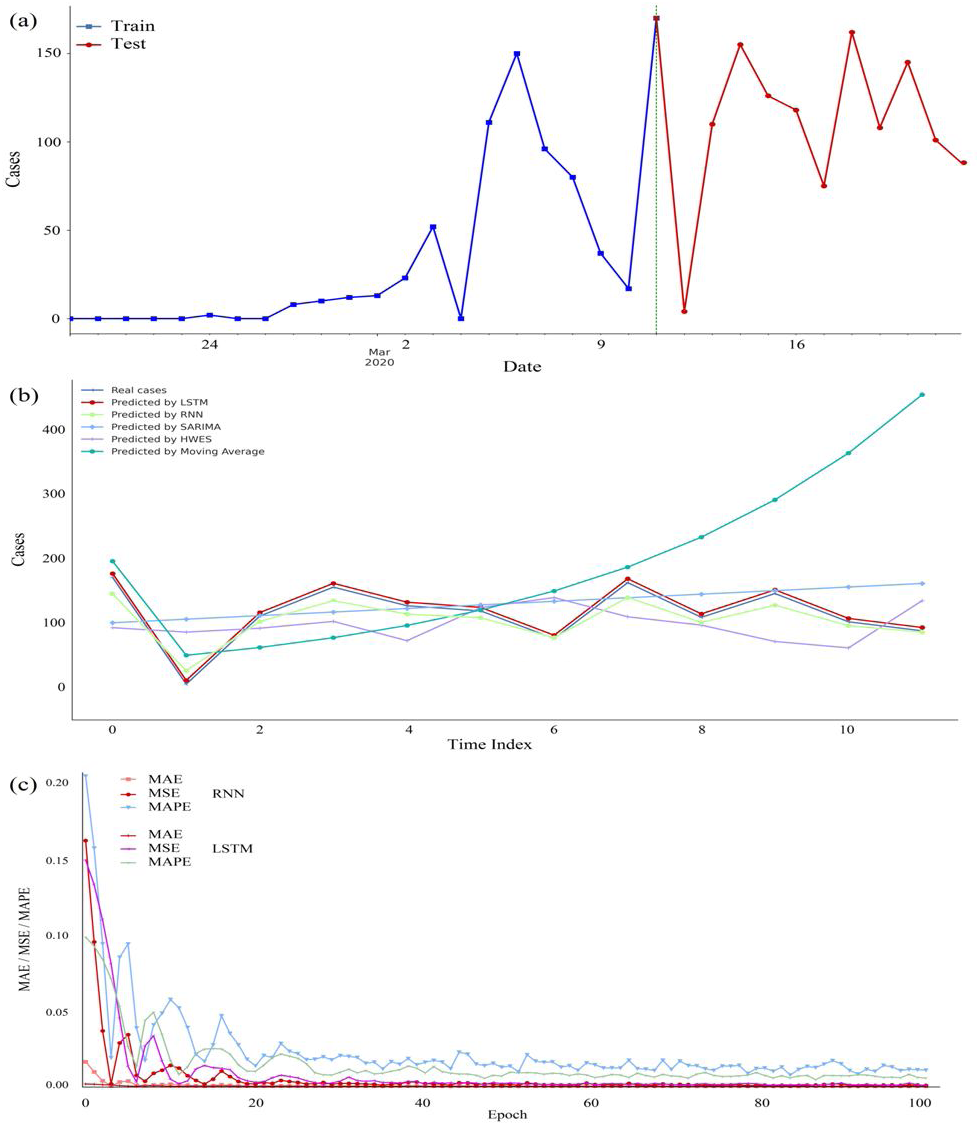
Infectious forecasting model for Isfahan province based on deep learning: (a) train and test days, (b) Forecasting and actual infected cases with different approaches (LSTM, RNN, SARIMA, HWES, and moving average), (c) Error models for RNN and LSTM

**Figure 6.**
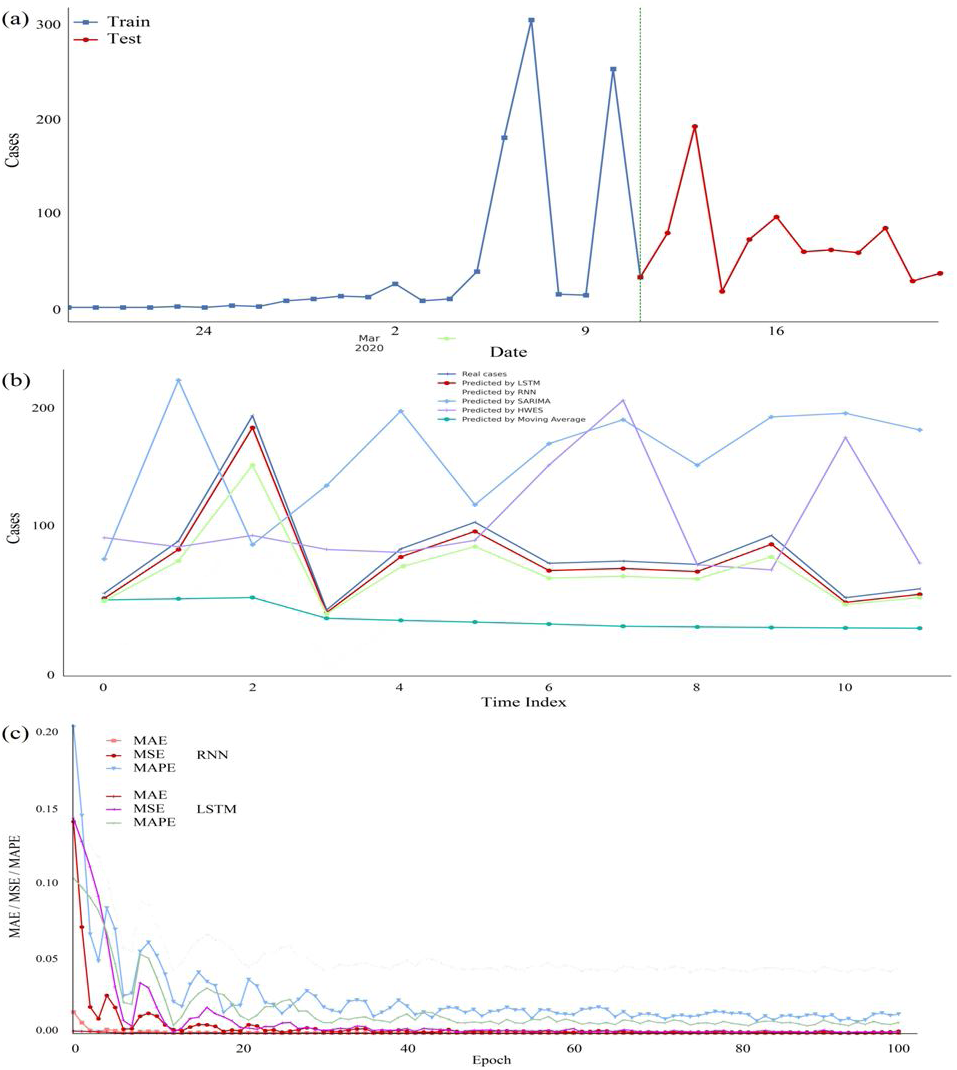
Infectious forecasting and actual infected for Mazandaran province based on deep learning: (a) Train and test days, (b) Forecasting and actual infected cases using different approach (LSTM, RNN, SARIMA, HWES and moving average), (c) error models for RNN and LSTM

**Figure 7.**
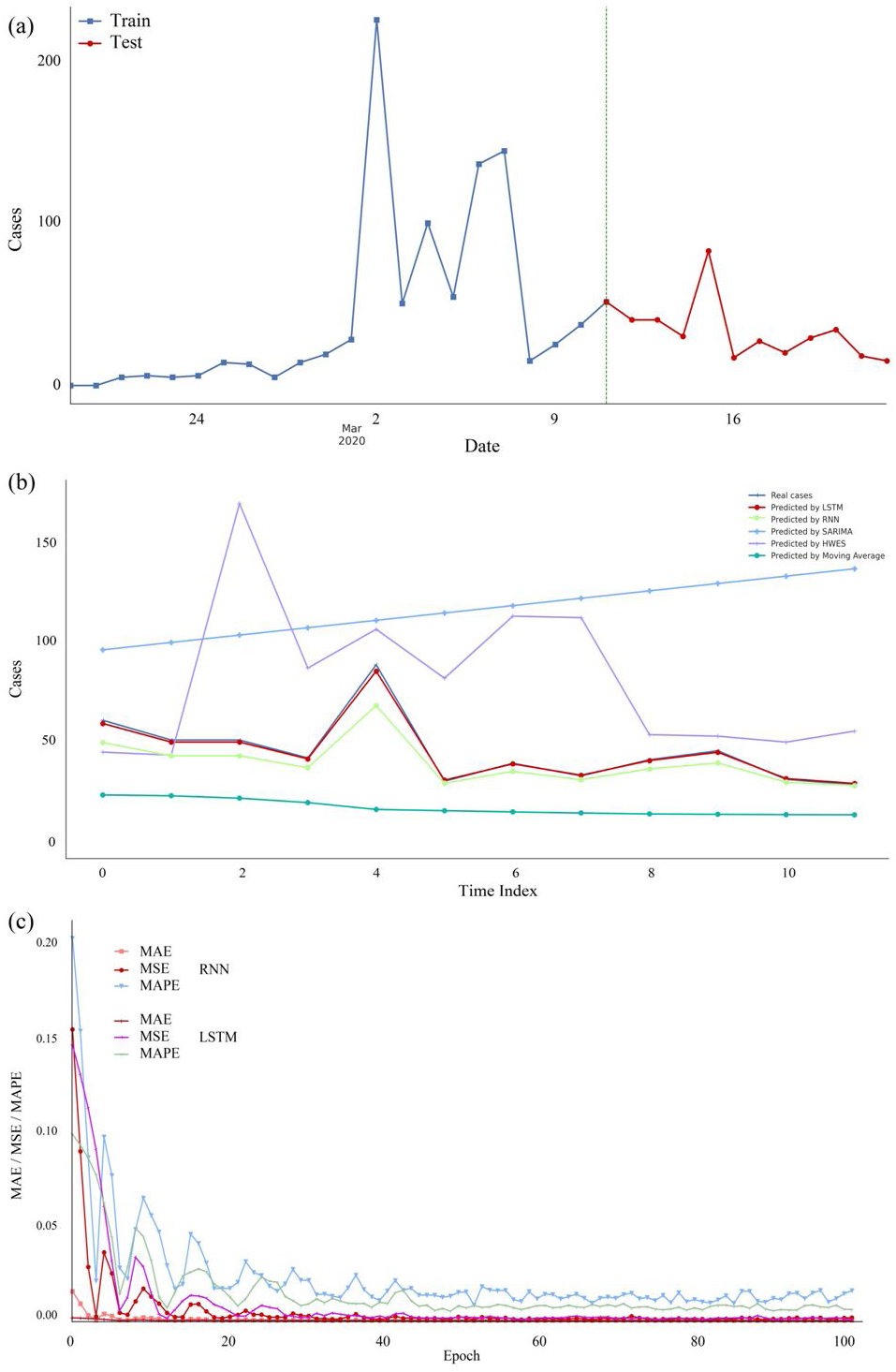
Infectious forecasting model for Qom province based on deep learning: (a) Train and test days, (b) Forecasting and actual infected cases using different approaches (LSTM, RNN, SARIMA, HWES, and moving average), (c) Error models for RNN and LSTM

**Figure 8.**
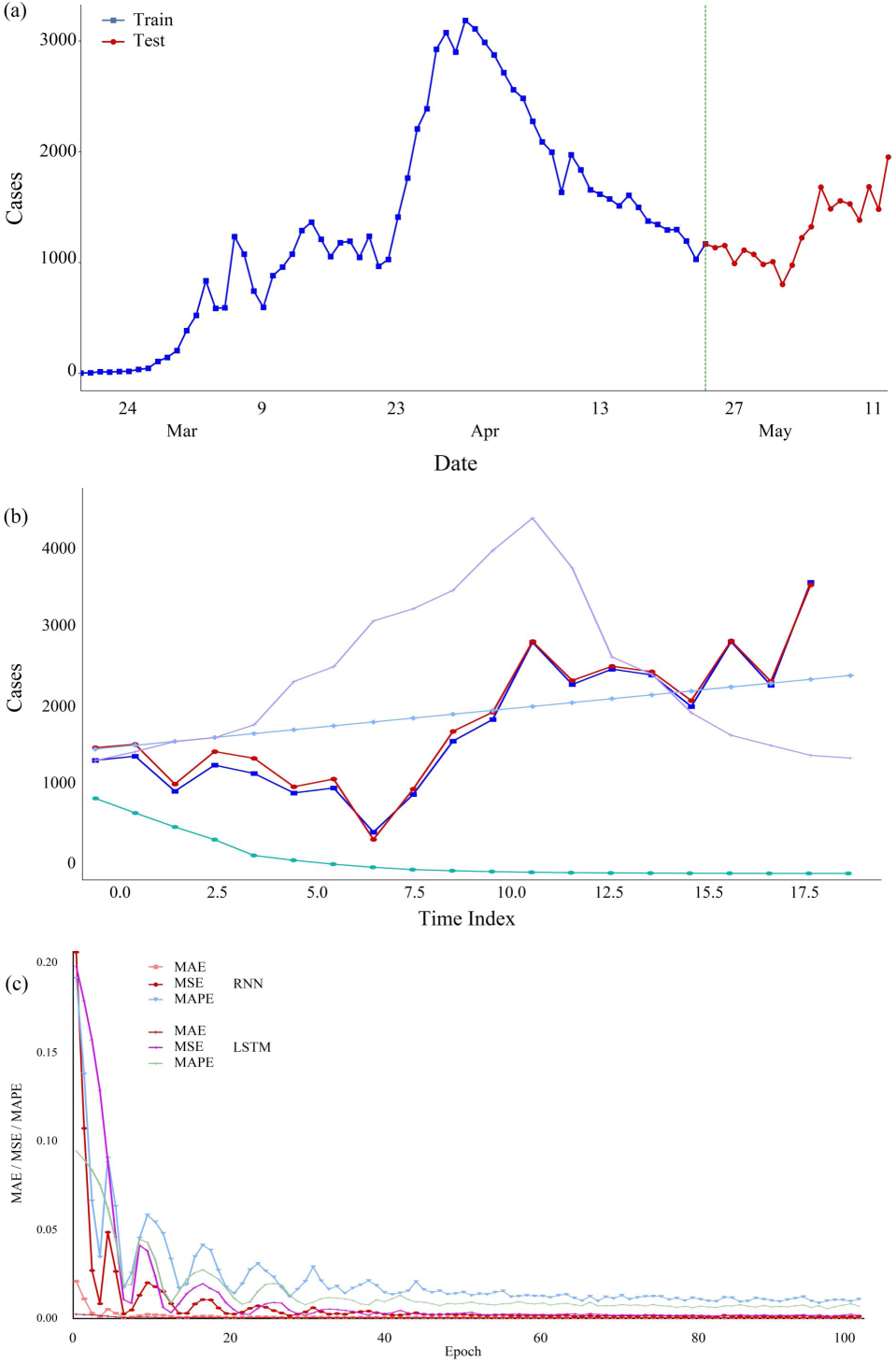
Infectious forecasting model for entire country based on deep learning: (a) Train and test days, (b) Forecasting and actual infected cases with different methods (LSTM, RNN, SARIMA, HWES, and moving average), (c) Error models for RNN and LSTM

**Figure 9.**
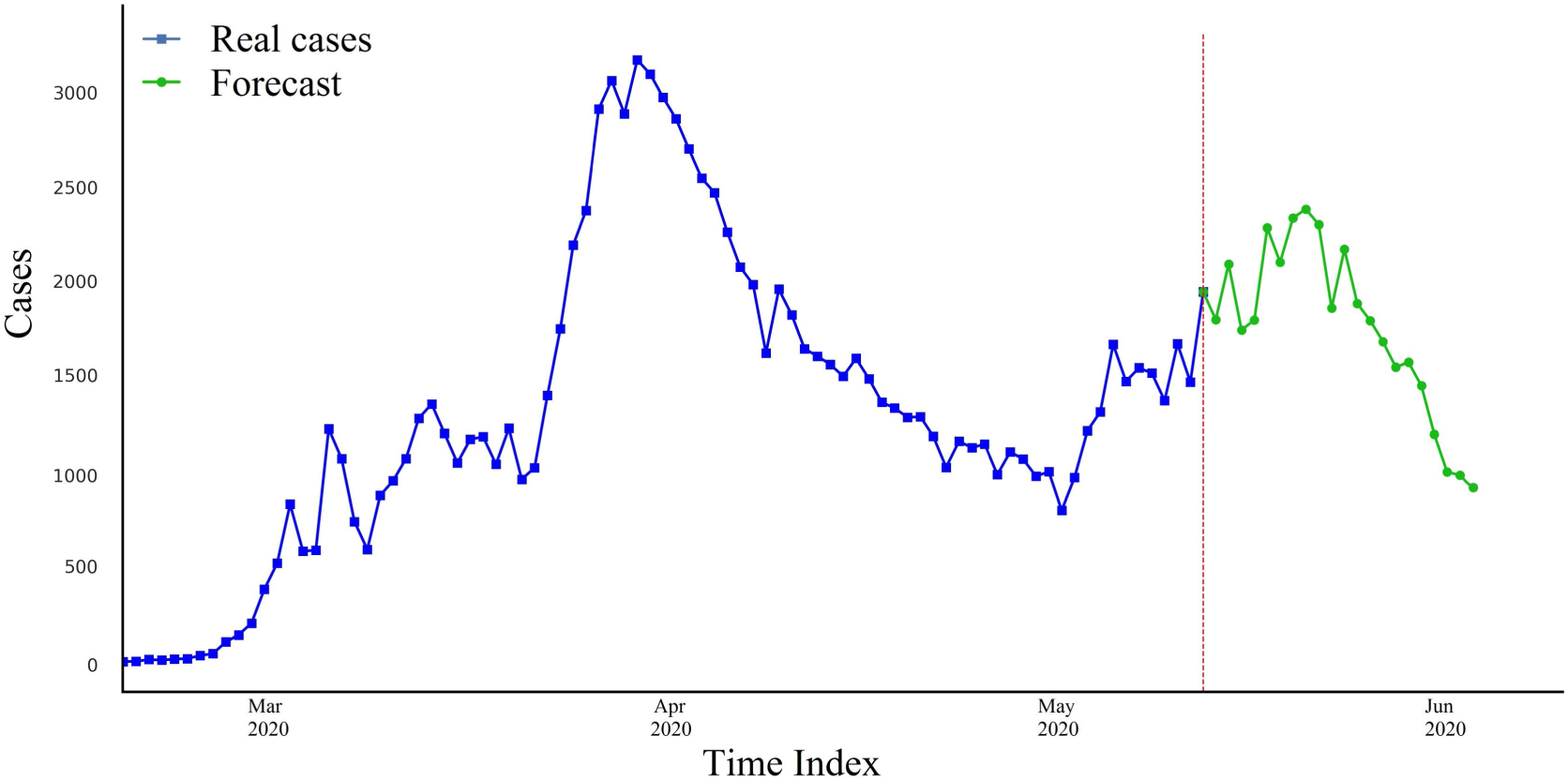
Forecasting and real cases variations in the next 21 days4.

## 4. Conclusion

The presented study attempted to use the LSTM neural network for time series modelling and forecasting of COVID-19 disease in Iran in the provincial and national levels. Provincial data collected from February 19 to March 22, 2020 and national data collected from February 19 to May 13, 2020 based on nationally recognised sources (e.g. Iran Ministry of Health and Medical Education; IRNA; ISNA). As methodology, the proposed model used a long short-term memory-based deep learning. We also compared the model to RNN, SARIMA, HWES, and moving averages approaches. Meanwhile, the MAE, MSE, and MAPE metrics were used to comparisons. Based on our experimental results, the LSTM model performed better than the other methods and gave less error values for infection development in Iran. According to the results, it can be stated that in the last days of the week, the infection cases will be decrease if the people follow current conditions and avoid crowded areas. According to the proposed model, the downward trend is likely to continue for up to three weeks (it should be noted that the rate of COVID-19 decay depends on various factors such as the person’s immune system, contact with others, underlying disease, exposure to polluted cities, non-compliance with social distances, travel to polluted cities and other factors). It seems that by the forecasting and evaluations, Iran has passed the first peak of the disease, but is possible to return the second disease peak in probable future.

## Data Availability

All data in the article

